# How, why and when are delayed (back-up) antibiotic prescriptions used in primary care? A realist review integrating concepts of uncertainty in healthcare

**DOI:** 10.1101/2023.11.07.23298228

**Authors:** Monsey Mcleod, Anne Campbell, Benedict Hayhoe, Aleksandra J. Borek, Sarah Tonkin-Crine, Michael V. Moore, Christopher C. Butler, A. Sarah Walker, Alison Holmes, Geoff Wong, the STEP-UP study team

**Affiliations:** National Institute for Health Research (NIHR) Health Protection Research Unit in Healthcare Associated Infections and Antimicrobial Resistance, Imperial College London, London, UK; NIHR Imperial Patient Safety Translational Research Centre, Imperial College London, London, UK; Department of Primary Care and Public Health, School of Public Health, Imperial College London, London, UK; NIHR Applied Research Collaboration Northwest London, London, UK; Nuffield Department of Primary Care Health Sciences, University of Oxford, Oxford, UK; NIHR Health Protection Research Unit in Healthcare Associated Infections and Antimicrobial Resistance, University of Oxford, Oxford, UK; Primary Care, Population Sciences and Medical Education, Faculty of Medicine, University of Southampton; NIHR Oxford Biomedical Research Centre, Oxford, UK; Nuffield Department of Medicine, University of Oxford, Oxford, UK

**Keywords:** antibiotics, delayed prescriptions, back-up prescriptions, deferred prescriptions, realist review, primary care, general practice antimicrobial resistance, antimicrobial stewardship, uncertainty, tolerance of risk

## Abstract

**Background:** Antimicrobial resistance is a global patient safety priority and inappropriate antimicrobial use is a key contributing factor. Trials have shown that delayed (back-up) antibiotic prescriptions (DP) are an effective and safe strategy for reducing unnecessary antibiotic use but its uptake is controversial.

**Methods:** We conducted a realist review (a literature review which goes beyond assessing whether an intervention works) to ask why, how, and in what contexts general practitioners (GPs) use DP. The review is focused on those who wish to use DP and not for those who are against using DP. We searched five electronic databases for relevant articles and included DP-related data from interviews with healthcare professionals in a related study. Data were analysed using a realist theory-driven approach – theorising which context(s) influenced (mechanisms) resultant outcome(s) (context-mechanism-outcome-configurations: CMOCs).

**Results:** Data were included from 76 articles and 41 interviews to develop a program theory comprising nine key and 56 related CMOCs. These explain the reasons for GPs’ tolerance of risk to different uncertainties—epistemological (knowledge-orientated); scientific (data-orientated); hermeneutic (interpretation-orientated); practical (structures/processes-orientated); technological (skills/software/equipment-orientated), and existential (world-view-orientated). These interact with GPs’ work environment, self-efficacy and perceived patient concordance to make using DP as a safety-net or social tool more or less likely, at a given time-point. Our program theory explains how DP can be used to mitigate some uncertainties but also provoke or exacerbate others.

**Conclusion:** This review explains how, why and in what contexts GPs are more or less likely to use DP, as well as various uncertainties GPs face which DP may mitigate or provoke. We recommend that efforts to plan and implement interventions to optimise antibiotic prescribing in primary care should consider these uncertainties and the contexts when DP may be (dis)preferred over other interventions to reduce antibiotic prescribing. We also recommend the following and have included example activities for: (i) reducing demand for immediate antibiotics; (ii) framing DP as an ‘active’ prescribing option; (iii) documenting the decision-making process around DP; and (iv) facilitating social and system support.

**SUMMARY BOX:**

- **What is already known on this topic** – Trials have shown that delayed (back-up) antibiotic prescriptions (DP) are an effective and safe strategy for reducing unnecessary antibiotic use but its variable uptake in primary care remains to be understood.
- **What this study adds** – This realist review provides a program theory to explain the complexity and interactivity of influencing factors on general practitioners’ (GPs) antibiotic prescribing decisions. It explains how GPs have a reasoned tolerance of risk to various uncertainties (beyond clinical uncertainty) which interact with GPs’ work environment, self-efficacy and perceived patient concordance to make using DP as a safety-net or a social tool more or less likely, at a given time-point. It applies nuanced concepts from the uncertainty literature - epistemological uncertainty (knowledge-orientated), scientific uncertainty (data-orientated), hermeneutic uncertainty (data interpretation-orientated), practical uncertainty (structures/processes-orientated), technological uncertainty (skills/software/equipment-orientated), and existential uncertainty (world-view and identity-orientated) - to better understand DP clinical decision-making.
- **How this study might affect research, practice or policy** – Policy makers, commissioners, and prescribers who would like to optimise antibiotic prescribing should become familiar with the varieties of uncertainties at play when GPs consult with patients and consider how these different uncertainties are mitigated and/or provoked when developing support interventions to optimise DP or implementation support.

## BACKGROUND

Drug-resistant infection associated with antimicrobial resistance (AMR) was attributed to 1.27 million deaths globally in 2019[1] and is a global patient safety priority.[1] Antimicrobial stewardship (AMS) promotes and monitors prudent use of antimicrobials, and is critical in healthcare to maximise patient benefit from immediate antibiotic treatment and to preserve future antimicrobial effectiveness by decelerating the development and spread of AMR.[2] Trials have demonstrated effectiveness of many AMS strategies within primary care.[3] One strategy, with evidence dating back to 1997[4] and advocated in national guidelines[5,6] is delayed (back-up) antibiotic prescriptions (DP). DP has shown to be effective for managing acute cough, acute sore throatClick or tap here to enter text., acute otitis media, sinusitis, and lower urinary tract infections.[7–20] DP is a prescription given to a patient (directly, or by post-dating, recontact, or collection[21]) with advice that antibiotics are not needed immediately and how to use the DP if symptoms worsen or do not improve after a certain time.[6,22,23]

Evidence suggests DP may reduce antibiotic use by patients[21,33] and may mitigate clinical uncertainty by providing a safety-net to minimise the risk of developing severe complications,[15,34–38] whilst maintaining patient satisfaction and not adversely impacting symptom severity or duration and reconsultation rates.[7–9,39] It is considered safe for most patients, including some in higher risk subgroups.[39] DP helps patients understand that antibiotics are not always needed, empowers patients in self-management, and reduces reconsultations.[40]

Those who do not favour DP, argue that the risk of harm from delayed antibiotics to an individual may outweigh potential benefits from reduced antibiotic use[41] and that DP confuses patients through mixed messages about antibiotic appropriateness.[28] Clinicians may be reluctant to transfer the responsibility of treatment decision-making to patients, believing it to unfairly burden the patient[28,37,42–47] or that the patient might use the antibiotic immediately or store for future use.(13) DP may also be less relevant in practices where reconsultation is the preferred safety-net[29,43] or in some rural and dispensing practices.[40]

Around a quarter of a century after the first evidence of DP as an effective AMS strategy,[4] a lack of reliable routine DP data hinders assessment of its uptake[7,24,25] and our understanding of what drives general practitioners’ (GPs) decision to use DPs or not is unclear. This study does not seek to resolve the controversy behind the pros and cons of DP use by GPs. Instead, the aim is to explain how, when and why it is used, therefore enabling the findings to inform design of DP implementation strategies in relevant infection pathways to support shared decision-making and enhance patient care.

## METHODS

We have conceptualised DP use by GPs as a complex phenomenon where usage (or not) is likely to be dependent on context. Unlike other forms of systematic reviews, which summarise the effectiveness of an intervention, we have conducted a realist review to understand how *context* influences the implementation of an intervention. A realist review asks how, when, why, and in what circumstances an intervention works.[51] Our realist review had six iterative overlapping stages[52] conducted in accordance with the Realist And Meta-narrative Evidence Syntheses: Evolving Standards (RAMESES).[53] This study is part of a wider research program (STEP-UP, https://www.expmedndm.ox.ac.uk/step-up). Our stages of the review process are set out in **Table 1** and **Box 1** highlights the additional concepts of uncertainty in healthcare literature that was also used to develop our program theory.

**Table 1.** Stages of realist review.

|  |
| --- |
| **Stage 1: Locating existing theories** |
| An initial programme theory was drafted based on the collective expertise of the project team (spanning general practice, pharmacy, infectious diseases, public health, health psychology, medical sociology and specialist realist methodology). The initial programme theory was iteratively refined following cycles of stages 2-6. We focused our review on GPs (or equivalent in different countries), due to differences in professional training compared with nursing, pharmacy and other prescribers and because GPs prescribe the majority of antimicrobials in primary care. |
| **Stage 2: Searching for evidence** |
| Systematic literature searches were carried out in Medline, EMBASE, Web of Science, Cochrane Database of Systematic Reviews, and PsycINFO using various search terms for delayed prescription (DP) (**Supplemental file 1**), last search February 2021). We also used interview data (full transcripts rather than published excerpts only)[54] and feedback from experts (**Supplemental file 2**) to strengthen our evidence base and to provide up-to-date insights into DP use in practice. |
| **Stage 3: Selecting articles** |
| All article titles and abstracts were independently screened (by at least two researchers) and selected full papers were reviewed by two researchers. Disagreements were resolved by consensus or via a third researcher. All research study designs and article types (empirical research, reviews, and perspectives) that provided a description of how DP was used or implemented in primary care settings were included. For non-research articles, only those that presented viewpoints or analysis of DP use in practice were included. We also included use of DP reported for any current infection for patients of all ages and excluded those used as rescue treatment for a potential future infection. |
| **Stage 4: Extracting and organising data** |
| We developed and applied internal processes to facilitate consistency among researchers for coding and analysis using a constant comparison approach[55] and organised our data in NVivo v.11 (QSR International). Six articles were first inductively coded by two researchers independently. Codes among researchers were then compared and refined, with similar codes grouped together into categories to create an early coding framework which was then re-applied to 20% of the remaining articles. A further iteration of the coding framework was produced as codes were again compared and refined. The final coding framework was applied to all articles. |
| **Stage 5: Synthesising evidence and drawing conclusions** |
| Stage 5 overlaps with stages 1, 4 and 6, and involved an inductive-retroductive-deductive analytic approach.[55–58] We used data and *inductive* coding framework (Stage 4) to identify prescribing outcome patterns. From this, we developed concepts and integrated these with the initial program theory (Stage 1), iteratively refining the concepts and our program theory as informed by data and substantive theory (Stage 6). Three researchers used *retroduction*[58] to infer explanations of the causal process for how a prescribing decision outcome pattern may occur, i.e. which context(s) influence (mechanisms) which in turn cause outcome(s) (context-mechanism-outcome-configurations: CMOCs) (**Supplemental file 3**). Explanatory CMOCs were used *deductively* in our analysis– i.e. used our data to confirm, refute and extensively refine the CMOCs until judged that they accounted for as much of our included data as possible. |
| **Stage 6: Searching for and engaging with substantive theories** |
| During refinement of our program theory, uncertainty was an important concept that needed to be understood.[59–65] We sought to further understand the role of uncertainty in relation to DP use.[15,34–38] From additional exploration of the ‘uncertainty’ literature (Box 1), we were able to refine a related concept of ‘tolerance of risk’, identified earlier in our program theory. Our final ‘tolerance of risk’ concept was defined as the GP’s emotional and/or cognitive, and subsequent behavioural, response to uncertainty about (i) whether antibiotics would be effective, necessary, beneficial or harmful; and (ii) how DP may mitigate or augment harms or benefits—to the patient or GP (e.g. medico-legal, reputational, trust). |

### Box 1. Concepts of uncertainty in healthcare literature which may be helpful to understanding DP use

#### Fears and hopes of uncertainty

- When uncertainty is present, clinicians will usually be guided by the widely held professional principle of non-maleficence (‘to do no harm’). This is typically associated with risk of adverse events (e.g. from treated or untreated infections) and negative emotions. This may manifest as fear and anxiety about missing diagnosis, feeling vulnerable to complaints or disciplinary action. Clinicians may usually be motivated to avoid, mitigate or prevent uncertainty as much as possible or practical[62,63] (e.g. by prescribing immediate antibiotics).
- Uncertainty also pertains to beneficence (‘to do good’) and to positive emotions (e.g. courage, confidence, curiosity) and hope for beneficial outcomes, e.g. that the patient learns that their illness may be self-managed without antibiotics. Clinicians may thus be motivated to ‘embrace’ uncertainty[66,67] when they realise that, paradoxically, some benefits are gained only when risks are taken.[68,69]

#### Early conceptualisations of uncertainty in healthcare

- Uncertainty in healthcare is commonly understood as ‘clinical uncertainty’—where medical knowledge, though dynamically changing with generation of new knowledge, is *always* limited, as is the mastery of this knowledge by any one individual doctor.[59,64,65]
- One aspect of this knowledge is information about the risk (probability) of a beneficial or adverse event (e.g. probability that 20% of patients will benefit or be harmed from treatment). Uncertainty may arise from ambiguity of this risk information regarding its reliability, credibility or adequacy (e.g. confidence interval of 10-30% probability that patients will benefit/be harmed with expert disagreement on benefits/harms, and insufficient scientific evidence). It may also arise from its complexity, when its features make it difficult to understand or apply (e.g. 20% probability of benefit/harm for a certain type of patient with/without a multiplicity of risk or causal factors and interpretive cues).[63]

#### Towards a more nuanced conceptualisation of uncertainty in healthcare

- Several types of uncertainty are already conceptualised in the wider healthcare literature,[62–64,70,71] and these may be important to understanding use of DP as an AMS strategy:

**- epistemological** (around knowledge)[70] and **scientific** (around data, diagnosis, prognosis, cause, and treatment)[62,63] (both of which may be akin to clinical uncertainty)
**- practical** (around structures and processes of care)[62,63]
**- technological** (around use of equipment or software)[70]
**- hermeneutic** (around interpreting data, e.g. test results or patients’ narratives)[70]
**existential** (around issues of personal identity, worldview or meaning,[63,64,70,71] from professional (what it means to be a ‘good’ doctor)[70] and patient[63,71] perspectives).

## RESULTS

We included 76 articles (**Supplemental file 4**), comprising 50 empirical studies, nine literature reviews and 17 commentaries. We also included excerpts from interviews with 22 healthcare professionals involved in local AMS implementation and 19 general practice professionals ( Supplemental file 2)

### Program theory: Influence of uncertainties and risk tolerance in context on DP use

Our program theory explains how different antibiotic decision-making contexts may shift during a primary care consultation and that DP decisions are influenced by two important concepts, namely the way GPs respond to different types of uncertainties and their tolerance of risk to these uncertainties.

Broadly, for GPs not ideologically averse to DP, we found that when GPs had a lower tolerance of risk but were uncertain if an immediate antibiotic prescription was needed, then they were more likely to use DP. In this situation, as scientific and epistemological uncertainty increased (see **Box 1** for types of uncertainties and **Supplemental file 5** for an illustration of the program theory), the GPs tended to increase DP use. However, the behavioural pattern above is influenced by: 1) perceived value of DP as a safety-net; 2) perceived patient concordance (i.e. perceived agreement after a collaboration to incorporate “the hopes, beliefs and actions of prescriber and recipient”[72] and GP self-efficacy to facilitate this; 3) perceived value of DP as a social tool; and 4) work environment (**Table 2**). Importantly, the context may change during the consultation as more information becomes apparent. Our program theory illustration (Error! Reference source not found.**Supplemental file 5)** represents snapshots of different antibiotic decision-making contexts, and how these may shift, from a nominal starting point (e.g. 61% immediate; 25% no antibiotic; 13% DP; for acute cough[7) during a primary care consultation.

**Table 2.** Program theory comprising four broad areas of influence encompassing nine context-mechanism-outcome-configurations (CMOCs) -key theories explaining general practitioners’ (GPs) decision-making for immediate antibiotic prescription, delayed prescription (DP), or no antibiotic. CMOCs 1.1 -9.10 provide further granularity on specific contexts and mechanisms that when present may increase or decrease use of DP.

| Key CMOCs (Context-Mechanism-Outcome-Configurations) |  | Related CMOCs |  |
| --- | --- | --- | --- |
|  |  | Likely to increase use of DP | Likely to decrease use of DP |
| **Value of DP as a safety-net** |  |  |  |
| 1. Antibiotic access | **CMOC 1:** When a GP is more uncertain about the need for an immediate antibiotic (C), they may be more likely to issue a DP (O) if they believe it provides a sufficient safety-net as patients can access antibiotics if needed (M)[15,32,34–38,40,44,54,74–78] | **CMOC 1.1:** Results of clinical prediction scores or diagnostic tests are intermediary or conflict with the GP's expectations, their uncertainty remains (C) [7,10,31,54,74,79–84]<br>**CMOC 1.2:** GP identifies constraints on patients' ability to access antibiotics (C) & wants to avoid burden of out-of-hours consultation or reconsultation for the patient (M) [7,31,35,40,42,44,45,47,54,74,78,80,82,85–88]<br>**CMOC 1.3:** GP uses their knowledge & interpretation of evidence / guidelines to support DP use (C) as this gives confidence to know when to use DP (M) or to justify approach to patient/others (M) [15,31,35,36,38,54]<br>**CMOC 1.4** GP perceives DP as an opportunity to reduce antibiotic use & contribute to wider AMR agenda (M) [11,13,15,31,34,40,44,47,76,78,85,89,90]<br>**Also evidenced with CMOC 2.1, 2.2, 3, 4.2, 5, 7, 8** | **CMOC 1.5:** Clinical scores/diagnostic tests results are high or low and GP manages conflicts with their expectations (C) [7,36,80,81,83,84,91]<br>**CMOC 1.6:** GP feels it either unsafe or unprofessional to transfer the decision to use antibiotics to the patient (M) [24,28,30,35,37,42–47,85,89,92,93]<br>**CMOC 1.7:** GP believes it safer to ask patients to reconsult (M); that patient may develop serious complications (M); or that DP delays patients seeking help for potentially serious illness (M); or wants to reduce medicolegal risk (M) [11,24,29,35,38,40,41,43–45,47,78,85,94]<br>**CMOC 1.8:** GP is not familiar with the patient (C) and believes there is less scope to follow-up patient (M) and is uncertain whether the patient will follow instructions for using DP (M)[35,37,44,47,74]<br>**CMOC 1.9:** GP believes DP should not be prescribed as it exposes patients to potential adverse drug effects and does not improve patient outcomes (M) [28,30,45,94] **Also evidenced with CMOC 3.3** |
| 2. Patients at high risk of developing complications | **CMOC 2:** If a GP is more uncertain about antibiotic need (C) & patient has co-morbidities or perceived as vulnerable or at risk of serious complications(C), they may issue immediate antibiotics (O) as they doubt DP is a sufficient safety-net (M) [15,35,38,40,44,54,85,89,95,96] | **CMOC 2.1:** GP interprets evidence/ guidelines to support use of DP (C) because this affords them a sufficient safety-net for a particular patient (M)[15,38]<br>**Also evidenced with CMOC 1.3** | **CMOC 2.2:** GP interprets evidence/guidelines that caution against the use of DP for certain patients (C) [35,75,97]<br>**Also evidenced with CMOC 1.6** |
| **Patient concordance and GP self-efficacy** |  |  |  |
| 3. Shared decision-making, ability to understand and use DP | **CMOC 3:** If GP perceives factors affecting a patient's ability to understand & follow DP instructions (C), GP is more or less likely to use DP (O) depending on their confidence that the patient will follow instructions as advised, intentionally or unintentionally (M) [98] | **CMOC 3.1:** GP perceives the patient to be sufficiently 'sensible' i.e. to understand, and agree with, the given reasons to delay (C) and trust them to delay filling the prescription (M) [40,42,44,45,47,54,74,85,89,99]<br>**CMOC 3.2:** GP perceives that the patient trusts them (C)[45,47]<br>**Also evidenced with CMOC 7.1** | **CMOC 3.3:** GP perceives the patient not to be educated or understand and follow instructions for DP as advised (with or without language barrier) (C) [18,20,40,44,45,54,74,85]<br>**CMOC 3.4:** GP perceives DP conveys a contradictory message about when an antibiotic is needed(C)[24,30,35,37,43,45,54,74,77,79]<br>**Also evidenced with CMOC 1.8, 4, 5.7** |
| 4. Feedback about DP use | **CMOC 4:** If GP lacks evidence (or it is insufficient, ambiguous, complex) of how patients use DP (C), they may not issue DP (O) as they are not confident patients will follow instructions, intentionally or unintentionally (M) [35,42,43,46,47,54,100] | **CMOC 4.1:** GP has improved audit and/or feedback on use (C) [16,35–37,44,54,75,101–103]<br>**CMOC 4.2:** GP controls access to the antibiotics through their choice of DP format/instructions (e.g. post-dating or asking patient to collect) (C) [35,40,44,54,80,85] | **CMOC 4.3:** GP perceives certain patients/groups to be more likely to not follow DP instructions as advised (e.g. working parents of young children who are struggling with childcare) (M)[42] |
| 5. GP self-efficacy in explaining the | **CMOC 5:** When a GP has high self-efficacy in explaining a DP (e.g. those with more experience) (C) they are more likely to issue a DP (O) because | **CMOC 5.1:** GP has a clear approach for communicating rationale & instruction for DP which may include providing reassurance to the | **CMOC 5.4:** GP uses communication skills/tools to discover patient does not actually want antibiotics (C) [41,46,47,75,103,112] |
| rationale & instructions for DP | they are more confident that it's a useful tool to manage their uncertainty and that the patient is able to understand why it is being given (M)[35,42,47] | patient, advice around self-care and/or symptom relief treatment (C) [15,17,21,24,33,38,74,76–78,85,99,100,103–107] see also[108]<br>**CMOC 5.2:** GP uses leaflets or other resources to help explain DP (C) [35,54,77,85,91,95,98,99,109]<br>**CMOC 5.3:** GP is provided with training about DP (C) [18,35,36,47,54,74,83,103,110,111] | **CMOC 5.5:** GP uses communication skills/tools to convincingly explain to the patient the rationale for not giving any antibiotics & provides self-care advice and reassurance (C) [11,35,37,54,77,94,105,106]<br>**CMOC 5.6:** GP worries that the patient may think them incompetent (C) or about reputational damage (M) [11,78,79,85,92]<br>**CMOC 5.7:** GP believes DP reinforces patients' erroneous beliefs of efficacy of antibiotics for self-limiting conditions (C) [28,29,37,43,54,94]<br>**CMOC 5.8:** GP uncertain about which DP format to use & how (C)[35,54] **Also evidenced with CMOCs 1.9, 3.3, 3.4** |
| **Value of DP as a social tool** |  |  |  |
| 6. Minimising conflict | **CMOC 6:** When a patient directly asks, or GP perceives them to expect antibiotics (C) & when GP is more certain that antibiotics aren't needed (C) GP may issue DP instead of no antibiotic (O) because they see it as a way to minimise conflict with the patient and/or patient dissatisfaction (M) [12,18,35,36,40,44,47,74,76,77,80,85,98,99,104] | **CMOC 6.1:** GP can give DPs in the most accessible format (C) [35,37,40,99] see also [21]<br>**CMOC 6.2:** GP uses DP as a compromise for patients who may have previously been given unnecessary antibiotics[11,35,37,80,88,112]<br>**CMOC 6.3:** GP wants to prevent patients accessing antibiotics another way (C/M) [35,44,45,54]<br>**Also evidenced in combination with CMOCs 1.4, 8.1-8.4, 9.2** | **CMOC 6.4:** DP format used in a GP's practice is barrier to prescription use (C) because they are concerned this to be perceived as untrusting by patients (M) [37,54,112] see also [21]<br>**CMOC 6.5:** GP/patient has preceding negative experience of DP(C)[96]<br>**CMOC 6.6:** GP/patient has preceding negative experience of not prescribing (or being prescribed) immediate antibiotics (C) [35,40,41,47,80,102] **Also evidenced with CMOCs 5.6, 5.7** |
| 7. Patient education & empowerment | **CMOC 7:** Across various contexts (C) a GP is more or less likely to issue a DP (O) because they believe it offers an opportunity to educate the patient that antibiotics are not always needed (M) or believe it beneficial to empower the patient to make illness management decisions (M)[11,12,14,19,37,38,40,42,44,45,47,74,75,85,102,113] | **CMOC 7.1:** GP perceives patient as receptive to learning about when antibiotics are not needed (C) or to having some control about when to use antibiotic (C)[18,35,42,44,75]<br>**CMOC 7.2:** GP believes that educating patient about DP can save GP/patient time (C/M)[4,15,19,47,76,78,88,96]<br>**CMOC 7.3:** GP believes DP contributes to a positive patient-practitioner relationship (C)[35,37,45,85,88,103]<br>**CMOC 7.4:** GP views DP as a 'psychological safety-net' for patients, validating and reassuring them their concerns were heard and addressed (C/M)[4,11,24,34,35,40,42,45–47,85,88,91,113]<br>**CMOC 7.5:** GP perceives certain patients/groups to be more likely to want to avoid antibiotics e.g. parents of young children (C)[44,96]<br>**Also evidenced with CMOCs 1, 3.1, 3.2** | **CMOC 7.6:** GP believes the patient may view DP as a signal of rejection of their symptoms (C/M)[91]<br>**Also evidenced with CMOC 1.6** |
| **Work environment** |  |  |  |
| 8. Workload & time pressures | **CMOC 8:** When a GP is constrained by high workload and/or time pressure (C) they may or may not issue a DP (O) depending on the extent to which they believe it to be the most efficient thing to do (M)[35,41,46,54,77,78] | **CMOC 8.1:** GP believes DP takes no extra time to explain (C/M)[34]<br>**CMOC 8.2:** GP believes DP saves time in other ways (C/M)[38,42]<br>**CMOC 8.3:** GP consults at certain times of day/week that drives them to complete consultations quickly (C)[54]<br>**CMOC 8.4:** GP wants to avoid the patient seeking out-of-hours care[74] (C) and /or workload for out-of-hours (C)[74]<br>**CMOC 8.5:** GP uses templates or shortcuts to resources(C)[54]<br>**Also evidenced with CMOC 1.2** | **CMOC 8.6:** GP believes issuing a DP takes more time to explain during the consultation (C)[35,46,47]<br>**CMOC 8.7:** GP believes that aspects of administering DP will create more work for the patient, GP or other staff (C)[35,40]<br>**CMOC 8.8:** GP uses a cumbersome electronic prescription transfer service (C)[54] |
| 9. Practice culture & structure | **CMOC 9:** When a GP experiences or perceives that DP is accepted & routinely used by their peers (C), they are more likely to issue a DP (O) because they are more confident that the patient will be | **CMOC 9.1:** And the practice or local guidelines consistently promotes DP for certain indications (C)[54,85,111]<br>**CMOC 9.2:** Practice consistently promotes DP format (C) [12,35,54,92,99] see also [21] | **CMOC 9.6** GP finds it hard to monitor their own or their practice's use of DP e.g. through coding of DP (C)[54]<br>**CMOC 9.7:** Where the practice has a policy of not prescribing antibiotics for certain infections (C)[35,37,54,92] |
|  | receptive to receiving a DP and to follow instructions for its use (M)[35,75,111,114] | **CMOC 9.3:** Monitoring performed & acted on by local or national systems to reduce immediate antibiotic prescriptions (C)[35,37,54]<br>**CMOC 9.4:** Visual cues in practice reminding GP of DP use (C) [101,115]<br>**CMOC 9.5:** Fee-for-service remuneration health-care model (C) with DP 'counting' to be remunerated (C) to maximise GP income (M)[28] | **CMOC 9.8:** GPs are aware of multiple guidelines/policies (local & national) with confusing or conflicting information (C)[35]<br>**CMOC 9.9:** Inconsistent DP use among prescribers (C) makes GP doubt how DP can be used optimally (M)[35]<br>**CMOC 9.10:** Fee-for-service remuneration health-care model (C) with reconsultation incentivised (C) to maximise GP income (M)[34,46,47] |

Within the wider program theory, we identified nine *key*[73] interacting context-mechanism-outcome configurations (CMOCs) that explain shifts in a decision between immediate, DP or no antibiotic (**Table 2**). Each *key* CMOC was further broken down into two or more *related* CMOCs (n=56) to provide further granularity on contexts and mechanisms that may increase or decrease the likelihood of DP use. We highlight examples associated with different types of uncertainty under each explanation (**Supplemental file 6**).

#### I. The value of DP as a safety-net

The safety-netting value of a DP is apparent when a GP is scientifically or epistemologically uncertain about whether the infection is viral, bacterial, self-limiting and/or whether a patient would deteriorate without timely antibiotics, and believes DP allows patients access to antibiotics if needed (CMOC:1). This is illustrated by an increased likelihood of using DP with increasing scientific and epistemological uncertainty. However, other present contexts (**Table 2**) may mitigate or provoke other uncertainties experienced by the GP and may influence their decision to use DP to safety-net. For example, when a GP identifies constraints on a patient’s ability to access antibiotics, this may provoke practical or technological uncertainty in addition to their scientific/epistemological uncertainty. This may be mitigated by the GP issuing a DP in an accessible way (CMOC:1.2) or, if their work environment allows, instead offering reconsultation to mitigate scientific uncertainty about illness progression and medico-legal concerns (CMOC:1.7). Another example is when a GP uses clinical scores or diagnostic tests: a clearly high or low score/test result may mitigate scientific uncertainty and lead to an immediate or no antibiotic decision (CMOC 1.5); an intermediary result may provoke hermeneutic uncertainty and lead to DP becoming preferred (CMOC:1.1). Moreover, all these contexts and uncertainties may be influenced by a GP’s existential uncertainty regarding their worldview of what makes a ‘good’ doctor: e.g., beliefs around the appropriateness of transferring antibiotic decision-making to patients (CMOC:1.6); or around the risks and benefits of giving patients potentially unnecessary medication (CMOC:1.9). For patients perceived at high-risk of serious complications, fear of missing a serious diagnosis is the main reason for using immediate antibiotics. DP use was influenced by how GPs’ scientific/epistemological uncertainty was addressed when they interpret evidence/guidelines as supporting DP for vulnerable patients as a safety-net (CMOC:2.1) or cautioning against (CMOC:2.2). Determining high-risk patients may differ among GPs, e.g. patient’s age was associated with different perceptions of risk by different GPs. This scientific/epistemological uncertainty may reflect their confidence or experience with managing perceived high-risk patients. **Table 2** also shows further contexts and mechanisms that influence GPs’ use of DP to safety-net.

#### II. Perceived patient concordance and GP self-efficacy

We found that GPs’ beliefs about a patient’s ability to understand and follow DP instructions (CMOC:3), receiving feedback about patients’ use of DP (CMOC:4), and GPs’ self-efficacy to explain DP (CMOC:5), all influence their confidence that they and their patients will reach a shared understanding of DP and agreement for its use, i.e. patient concordance. When perceived patient concordance is low, e.g. when a GP believes a patient to have a lower level of education (CMOC:3.3) or does not know the patient sufficiently (e.g. when working out-of-hours) to assess whether they will use DP as intended (linked CMOC:1.8), then GPs may use DP less. Different types of uncertainty, arising from different contexts, may contribute to a GP’s perception of potential patient concordance and DP may mitigate (CMOCs:4.1-4.2, 5.1-5.3), provoke or exacerbate (CMOCs:3.4, 4.3, 5.6-5.8) these uncertainties. For example, existential uncertainty around patient concordance may be mitigated with attention to scientific uncertainty and improved data on how individual patients, or patients in general, use DP (CMOC:4.1). If perceived patient concordance is due to practical or technological uncertainty about how the patient will access the DP, then this may be improved by providing the GP with information about different DP formats which may facilitate/hinder access (CMOC:4.2). Practical and technological uncertainty may be further addressed by improving GPs’ self-efficacy in explaining the rationale and instruction for DP (CMOCs:5.1-5.3), and that antibiotics are not needed (CMOC:5.5), or to discover that the patient does not want antibiotics (CMOC:5.4).

#### III. Using DP as a social tool

In situations of less scientific/epistemological uncertainty and where there is perceived or actual patient demand for antibiotics, other uncertainties may predominate. Rather than using DP as a safety-net, some GPs used DP more as a social tool to address practical and existential uncertainty around their relationship with the patient, including minimising conflict (CMOC:6); educating and empowering patients to make illness management decisions (CMOC:7); and reassuring them their concerns were heard (CMOC:7.4). GPs may want to provide a DP which minimises patients accessing antibiotics another way (CMOC:6.3) and choose a DP format to suit them or their patient (CMOC:6.1) rather than one which patients perceive as obstructive or paternalistic (CMOC:6.4). GPs may believe DP acts as a reassuring ‘psychological safety-net’ for patients (CMOC:7.4), especially useful for those patients wanting to avoid antibiotics (CMOC:7.5). Practical and existential uncertainty is also relevant when GPs perceive patients as receptive to self-management (CMOC:7.1) and that educating them about DP can save GP and patient time by reducing future care-seeking behaviour for similar illnesses (CMOC:7.2). **Table 2** shows additional contexts and mechanisms (operating separately or together) that make using DP as a social tool more likely (CMOCs:6.1-6.3,7.1-7.5, 1, 1.4, 3.1-3.2, 8.1-8.4, 9.2) or less likely (CMOCs:6.4-6.6,7.6, 1.6).

#### IV. DP and the work environment

Two key work environment contexts influenced a GP’s prescribing decision: workload and practice culture. They found practical and technological uncertainty relevant when GPs have a high workload and feel under time pressure and want to take the most efficient approach for them or their patient (CMOC:8). If they perceive DP to take little additional time to explain (CMOC:8.1), or administer (CMOC:8.5), or to save time in the future (CMOC:8.2), they may use DP. Conversely, if GPs perceive DP to be more time-consuming than prescribing immediate antibiotics—either by explaining (CMOC:8.6) or administering DP (CMOCs:8.7-8.8), they may use DP less. Practical uncertainty here is also influenced by when the consultation occurs, e.g., Friday afternoon consultations, public holidays, or before an event important to the patient, may mean GPs use DP to enable patient timely access to antibiotics (CMOCs:8.3, 1.2) or to avoid seeking out-of-hours care (CMOC:8.4). Practical and technological uncertainty also depends on how the practice culture and structure supports routine DP use by peers (CMOC:9). Additional contexts or mechanisms here may be pertinent to scientific/epistemological uncertainty, e.g., incorporating (or not) DP use into local guidelines and the extent to which this is perceived as confusing (CMOCs:9.1, 9.7-9.9), or whether peer monitoring of antibiotic prescribing occurs (CMOCs:9.3, 9.6). This includes contexts/mechanisms that may attend to practical uncertainty by making practice approaches and beliefs more visible such as being clear about the variety of DP formats that GPs can provide to suit them or their patient (CMOC:9.2) and reminder cues (e.g., electronic alerts) to GPs about DP (CMOC:9.4). Practical uncertainty may also be important depending on the remuneration healthcare model in place (CMOCs:9.5, 9.10).

## DISCUSSION

Our program theory explains how, when and why GPs use DP in different consultation contexts. It posits that GPs have a reasoned tolerance of risk to a range of uncertainties: epistemological (knowledge-orientated); scientific (data-orientated); hermeneutic (data interpretation-orientated); practical (structures/processes-orientated); technological (skills/software/equipment-orientated), and existential (world-view and identity-orientated) (**Box 1**). These uncertainties interact (individually or together) with GPs’ work environment, perceived patient concordance and self-efficacy to influence the likelihood of issuing DP as a safety-net or social tool at a given time-point, which may shift as the consultation progresses. Our program theory corroborates and advances knowledge around the complex interacting manifold clinical, social, and moral factors that play a role in a GP’s decision to prescribe antibiotics.[40,41,44,54,116–118]

### DP may mitigate, provoke or exacerbate various uncertainties

We propose that use of DP may mitigate, provoke or exacerbate various uncertainties, beyond that typically understood as ‘clinical uncertainty’,[59,119] arising from different contexts in primary care consultations (**Table 2**). For example, although diagnostic tests are often perceived to help reduce clinical uncertainty[120,121] (more nuancedly, scientific/epistemological uncertainty), other literature (pre-COVID-19) suggests that some GPs consider clinical uncertainty rare[40] and interventions to address it thus less relevant or that such technologies may instead lead to more uncertainty over the meaning of results.[40,65,116] Wider literature identifies this uncertainty as hermeneutic uncertainty.[70] We propose hermeneutic uncertainty occurs when GPs have difficulty interpreting intermediate diagnostic/clinical score results; when results conflict with their clinical judgement; when there is uncertainty around a patient’s level of risk for developing serious complications; and when interpreting patients’ narratives to determine their expectations for antibiotics. We found that use of DP to safety-net may mitigate some uncertainties – hermeneutic, practical, technological, and existential – but, and importantly, it may also provoke them. For example, DP may mitigate practical uncertainty around how patients will access antibiotics if needed but provoke a different type of practical/technological uncertainty around which DP format to use and how.[35]

From an existential uncertainty perspective,[63,64,70,71] the safety or professionalism of transferring antibiotic decision-making responsibility to patients sits within moral philosophy arguments for GPs to consider patients and public outside the immediate consultation and future generations of patients in their antibiotic decision-making.[118] This view of what it means to be a ‘good’ doctor may be important not only for how GPs view their own professional role[40] but also, we show, GPs’ concern about whether patients may view them as ‘incompetent’. It is also relevant within the ‘too much medicine’ paradigm[122,123] where GPs, usually guided by principles of ‘doing good’ and/or ‘doing no harm’,[26,27] may be uncertain around the competing pragmatic benefits of DP and the ideal of no antibiotic, with some believing that DP may be a distraction in achieving this ideal. For example, using DP to help patients pragmatically self-manage illness may sit uneasy with those who believe this is better achieved without using DP[28] and who worry about counterproductive ‘harms’, e.g. iatrogenic effects of unnecessary antibiotics or unintentionally increasing antibiotic use.

### Impact of heuristics on DP perceptions and use

We have described how different aspects of the work environment influence GPs’ use of DP and time pressure (amongst others) was a key factor in their decision-making. To help overcome time pressure, some GPs used heuristics to make efficiency-thoroughness trade-offs (ETTO).[124] Heuristics, normal and necessary mental shortcuts to lessen cognitive load, involves seeing aspects of current situations similarly to previously experienced situations and where decision responses are transferred from the previous situation to the present. Example heuristics from our findings include, ‘my explanation has/hasn’t worked before, so it will/won’t work now’ and ‘it seems like the patient is (in)sufficiently “sensible” to understand DP, so DP probably isn’t/is appropriate’.[124] Although this transfer is clearly valuable as it uses previous learning to avoid assessing everything from scratch, it is also potentially problematic with the risk of negative transfer, where learned responses are inappropriate.[124] For example, negative transfer from heuristics around ‘clinical uncertainty’ can lead to action bias, anticipated regret, and risk aversion—all of which, in the context of antibiotic prescribing, tend behaviour towards action rather than inaction and overprescribing immediate antibiotics ‘just-in-case’ to prevent potential harm to patients.[125] From this perspective, DP is considered to be a form of inaction by some GPs and looked upon as a relatively high risk strategy compared with immediate antibiotic. For example, some GPs viewed DP as a type of inaction if the DP format hinders patient access to antibiotics.[41]

Some heuristics around DP are also related to human factors, i.e. “interactions among humans and other elements of a system”[126–128], including how DP is operationalised in different practice settings. Despite clinical trials and studies involving DP scripts, formats, and communication support tools,[21,99] uncertainty remains in clinical practice for how GPs can use DP and how the prescribing system and work environment supports or hinders this decision option. This includes misperceptions such as DP being limited to post-dating prescriptions and that it is not possible to issue DP via electronic transfer to community pharmacies in some health care systems.

To help address the challenges of heuristics, four psycho-social strategies have been proposed by Tarrant and Krockow,[125] derived from a range of health literatures, to mitigate antibiotic overprescribing: 1. Strategic framing of treatment options (e.g. frame immediate antibiotics as ‘low-value’ in some circumstances and DP as an ‘active’ option that provide more optimal benefit vs risk for individual patients); 2. Substitution – replacing an undesired behaviour with an alternative (e.g. provide evidence and implement guidance to support substitution of immediate antibiotic with DP for specific infections and situations); 3. Documentation – enabling and encouraging prescribers to document decision-making process and discussions with the patient to mitigate against accusation of negligence; 4. Social support - this recognises that antibiotic decision-making by an individual prescriber is often influenced by their interactions with colleagues and expert advisors as well as their own knowledge and experience. Interventions that enhance social support through senior or peer review or feedback from patients can therefore help prescribers to fine-tune their tolerance of uncertainty and discourage defensive medicine.[125]

### Implications for practice

For policy makers, implementation teams and researchers, **Table 2** could be used as a resource for reviewing why DP is working or not for certain patient cohorts, situations or settings to identify what potential interacting mechanisms are at play and what type(s) of uncertainty may be targeted for intervention.

In practice and as evidenced in this review, DP is often seen as a compromise to no antibiotics (and not just as an alternative to immediate antibiotics). This then has the potential unintended consequence of driving up antibiotic use. To mitigate this, we suggest efforts to support DP be explicitly presented as an alternative to immediate antibiotics i.e. when a GP is uncertain about the benefits of immediate antibiotic and DP provides an appropriate safety-netting tool (rather than primarily as a social tool). DP should only be used for certain infections where there is evidence of benefits for specific patient groups/situations and risks of harm can be minimised e.g. through additional safety-netting advice. Ultimately, the goal is to improve confidence, by both prescribers and patients, in how certain infections can be effectively and safely managed without antibiotics, supported by self-care advice. Our recommendations are therefore to consider how DP aligns with no antibiotic options and self-care advice from a broader infection management perspective, and we have framed these using the psycho-social strategies by Tarrant and Krockov (**Table 3**).

**Table 3.** Recommendations.

| Recommendation | Example tasks | Links to program theory |
| --- | --- | --- |
| **Strategic framing of infection treatment approaches** |  |  |
| **1. Reduce demand for immediate antibiotic.** For example, frame immediate antibiotics as ‘low-value’ for some patients with certain infections where there is relatively low patient benefit vs risk of antibiotic side effects. | i. Co-create with patient/public, and prescriber stakeholders, and carry out user testing of different ways to communicate evidence-based situations when immediate antibiotics offer ‘low-value’ for patients i.e. unlikely to have much benefit and more likely to have negative consequences for the individual such as side effects, practical inconvenience and cost etc. This includes information that dispels inaccurate preconceptions about how and when DP can be used and how such key messages can be routinely reinforced at practice/clinic and regional level (e.g. to support practice culture) and national level (e.g. to align prescriber and public-facing DP use options). Activities/tips (including use of patient leaflet) to help prescribers discuss DP and no antibiotic options with patients to enhance patient understanding of infection progression, self-management options and safety-netting.<br>ii. Identify where, in the patient health-seeking process, the low-value of antibiotics for some situations can be communicated and DP can be explained to better prepare patients to engage in shared decision-making. This includes communication about self-care, illness duration of different infections, side-effects of antibiotics and safety-netting advice.<br>iii. Explore how different uncertainties impact patient/public expectations for antibiotics.<br>iv. Investigate potential unintended consequences of strategic framing of infection treatment options and take action(s) to mitigate against negative consequences. | See key and related CMOCs in:<br>CMOC 1: Antibiotic access<br>CMOC 3: Shared decision-making & ability to understand DP & follow instructions<br>CMOC 6: Minimising conflict<br>CMOC 7: Patient education & empowerment<br>CMOC 8: Workload & time pressures<br>CMOC 9: Practice culture & structure |
| **Substitution** |  |  |
| **2. DP as an ‘active’ alternative to immediate antibiotic.** Make more explicit in national and local guidelines to emphasise ‘low value’ of immediate antibiotics in some circumstances and when the ‘active’ alternatives of DP, no antibiotic and self-care options should be used. | i. User testing of different ways to communicate to prescribers that DP is an evidence-based intervention and can be used effectively and safely as an alternative to immediate antibiotic. This includes providing an overview of current DP evidence (including how DP can contribute to positive patient-practitioner relationship, providing a ‘psychological safety-net’ for patients). Include explicit evidence summary to support interpretation of DP or no antibiotic for specific infection pathways and situations, and include guidance on how point-of-care tests and clinical scores may help to support shared decision-making and uncertainty e.g. for high-risk patient groups.<br>ii. Explore which patients under what circumstances may benefit most from use of DP in different locations i.e. general practices/clinics providing services to patients in rural areas may have different accessibility issues compared to those in urban areas and DP may be more or less appropriate depending on how the practice/clinic is set up.<br>iii. Promote awareness of different formats of DP and encourage prescribers to test and adopt format(s) that are suitable for their patient population within their practices/clinics, locally and/or regionally.<br>iv. Explore how to provide and implement consistent messaging and practical guidance for DP use in multiple communication channels such as guidelines, webinars, training materials etc. | See key and related CMOCs in:<br>CMOC 1: Antibiotic Access<br>CMOC 2: Patients at high risk of developing complications<br>CMOC 3: Shared decision-making & ability to understand DP & follow instructions for DP use<br>CMOC 5: GPs self-efficacy in explaining the rationale & instructions for DP<br>CMOC 6: Minimising conflict<br>CMOC 7: Patient education & empowerment<br>CMOC 9: Practice culture & structure |
|  | v. Refine/develop a communication and skills assessment tool and training which frame DP, no antibiotic and self-care as 'active' alternatives to an immediate antibiotic pathway. Embed tool and training into established providers e.g. national professional bodies and medical schools.<br>vi. Incorporate guidance on diagnostic tests and clinical scores to inform decision to use immediate, DP or no antibiotic. This includes guidance on documentation and coding of these tools and the prescribing outcome to enable prescribing behaviour to be reviewed and inform potential future quality improvement. |  |
| **Documentation** |  |  |
| 3. Capture (and feedback) data on infection consultations, prescribing decision, and use of DP | i. Use existing data where available on documented infection consultations, prescribing decisions i.e. immediate antibiotic, DP or no antibiotic, and format of DP provided etc. Data should be monitored for trends, identify potential good practices and areas for targeted improvement. Provide feedback to prescribers within the practice/clinic organisation to support<br>ii. Where (i) is limited or not available, carry out work to enhance documentation and coding pathways and/or develop robust processes to facilitate regular documentation and coding or infection consultations and prescribing decisions.<br>iii. Data should be captured at different levels to allow benchmarking e.g. at prescriber, practice, organisation, regional and national levels.<br>iv. Explore ways in which patient outcomes data (process and clinical) can be captured and linked with primary care infection consultations. | See key and related CMOCs in:<br>CMOC 4: Feedback about DP use<br>CMOC 9: Practice culture & structure |
| **Social and system support** |  |  |
| 4. Facilitate a practice culture for discussing and reviewing DP, no antibiotic and self-care options to enhance their use for relevant clinical situations (including heuristics, changes in workflow, use of technology) | i. Support prescribers to share their experiences of how DP, no antibiotic and self-care can be explained with minimal or no additional time in different scenarios (e.g. different patient groups, in/out-of-hours, remote consultations, interpretation of guidelines)<br>ii. Co-create and test ways in which technologies (such as electronic prescribing and remote consultations) may be optimised to facilitate appropriate DP use and capture DP-related data with minimum burden on prescribers (e.g. streamline coding workflow in electronic prescribing system)<br>iii. Regularly review, feedback and discuss impact of DP on consultation process, quality and safety, and agree actions for further improvement to enhance patient care and safety<br>i. Develop and/or encourage processes to facilitate patient feedback post-consultation on outcomes such as use of DP, duration of symptoms, re-consultation, hospital admission etc. Explore how post-consultation follow-up workload data and related outcomes may be captured.<br>ii. Explore approaches for optimising patient experience in the infection pathway including enhancing documented communication on previous infection diagnoses and treatments, support continuity of patient care where appropriate, facilitating consistent messages from prescribers on infection management and minimising conflict etc. | See key and related CMOCs in:<br>CMOC 1:Antibiotic access<br>CMOC 3: Shared decision-making & ability to understand & follow instructions for DP<br>CMOC 6: Minimising conflict<br>CMOC 7: Patient education & empowerment<br>CMOC 8: Workload & time pressures<br>CMOC 9: Practice culture & structure |

### Strengths and limitations

A key strength of this review is incorporation of more nuanced types of uncertainty, beyond clinical uncertainty, on how and why GPs use DP. By using a systematic theory-driven realist synthesis of the evidence, we have been able to identify mechanisms likely to be applicable across different types of interventions to facilitate appropriate DP use. Rather than be prescriptive about when DP should or should not be used over other interventions, our program theory recognises the complexity of shared decision-making and sheds light on what range of contextual factors tend to enable safe and effective DP use. It also highlights how each of these factors may potentially be addressed and that a multi-faceted approach to reducing different types of uncertainty in different settings is required.

Other strengths are using different types and sources of evidence, including empirical data and demonstrating face validity through collaboration with a multi-professional team of researchers, clinicians and subject-matter experts. Unlike conventional systematic literature reviews, a realist review generates a program theory which, if sufficient evidence was analysed appropriately, should withstand the test of time and subsequent evidence would more likely reinforce than dispute the core constructs. However, we mostly identified and used evidence from high-income countries and focused our review on the perspective of GPs and not on patients, or other prescribers, which may limit some transferability. All the evidence included in the review was collected by February 2021 and none referred to the COVID-19 pandemic; however, we discuss below the potential implications of our program theory in the context of a pandemic and increased use of remote consultations.

### Future Research

The COVID-19 pandemic has led to significant changes in health service delivery.[129,130] The need to reduce disease spread has meant minimising face-to-face appointments and a dramatic increase in remote-consulting along with reduced ability to examine patients.[129–132] It has also meant various changes in health-seeking behaviour, including reported delays in health-seeking advice by patients.[130] Changing COVID-19 testing requirements, healthcare access and provision challenges due to mandated total triage models have further complicated the consultation process.[129,130] Consequently, actual or perceived demand (and need) for antibiotics from those who do seek healthcare advice may have been higher, e.g. from individuals suffering symptoms longer. Post-pandemic antibiotic prescribing data are now emerging; these should be examined for longer lasting effects of the pandemic on prescribing habits or changing public/patient beliefs about antibiotics.

Here we consider how our program theory can be applied in the context of the COVID-19 pandemic which may have substantially increased various uncertainties. For example, GPs, who would normally see most patients face-to-face, may have felt uncomfortable assessing patients remotely, experiencing increased scientific uncertainty by not being able to physically examine patients or use diagnostic tests in the GP surgery. This may have been confounded early in the pandemic with practical and technological uncertainty when many were less experienced in remote consulting and it was unclear whether issuing a DP was practically or technologically possible. One recent interview study with GPs concurs, revealing mixed views on perceived DP usefulness and actual use since the pandemic.[130] Since the COVID-19 pandemic, widespread self-testing has become standard for many patients and may facilitate uptake and acceptance of other point-of-care testing by patients in the future.

Further research is needed to understand and improve how different uncertainties are self-managed by patients. Where a GP or other healthcare professional (e.g. nurse practitioner, community pharmacy staff) is involved, research is needed to better understand how uncertainties are communicated to patients, and how some of these may be mitigated or provoked by DP. This includes improving how patients, GPs and other healthcare professionals may develop better DP explanations on when and how to use it. Further research may also be carried to support implementation of our recommendations in **Table 3**.

## CONCLUSION

This review provides practical insights for considering how various uncertainties influence antibiotic decision-making and DP-related behaviour. We show how different uncertainties (beyond clinical uncertainty)—epistemological, scientific, hermeneutic, practical, technological, existential— interact with GPs’ work environment, perceived patient concordance and GPs’ self-efficacy to influence use of DP to safety-net or as a social tool in primary care. Efforts by policy makers, commissioners, and prescribers to optimise antibiotics can use our program theory to help identify, design and implement interventions to address these uncertainties. We also recommend activities to: 1) reduce demand for immediate antibiotics by reframing infection management strategy options and emphasising clinical situations where there is relatively little or no benefit from immediate antibiotics; 2) reinforce DP as an ‘active’ prescribing alternative to immediate antibiotic; 3) document the decision-making process around DP; and 4) social and system support to facilitate a practice culture for using and improving infection management (including use of DP).

## SUPPLEMENTAL FILES

Supplemental file 1 – Primary literature search strategy

Supplemental file 2 – Document characteristics, source, use and nature of evidence Supplemental file 3 – Categorising contexts, mechanisms and outcomes Supplemental file 4 – Literature selection, inclusion and exclusion

Supplemental file 5 – Program theory on delayed prescription decision-making Supplemental file 6 – Summary program theory CMOCs and uncertainty types

## AUTHOR CONTRIBUTION STATEMENT

All authors contributed to the design of the study and its development. MMc and BH developed the search strategy with support from a specialist librarian. Articles were screened, data were extracted, and early themes developed by MMc, BH, AC and AB. The initial program theory was developed by MMc and BH, presented and discussed with wider STEP-UP team, then iteratively refined with all authors and substantial input from AC and GW. MMc and AC conducted the principal analysis and GW, BH, AB, STC contributed to the interpretation and presentation of results to the wider STEP-UP team. Iterative refinement and preparation of draft manuscripts were prepared by MMc and AC with contributions from GW. All authors provided critical feedback and helped shape the research, analysis and manuscript. All authors have read and agreed to the final version of the manuscript.

## Data Availability

All data produced in the present study are available upon reasonable request to the authors

## ACKNOWLEDGMENTS

This review is part of the work of the STEP-UP team (https://www. expmedndm.ox.ac.uk/step-up/step-up) comprising: Philip E Anyanwu, Aleksandra Borek, Nicole Bright, James Buchanan, Christopher Butler, Anne Campbell, Ceire Costelloe, Benedict Hayhoe, Alison Holmes, Susan Hopkins, Azeem Majeed, Monsey McLeod, Michael V Moore, Liz Morrell, Koen B Pouwels, Julie V Robotham, Laurence S J Roope, Sarah Tonkin-Crine, A. Sarah Walker, Sarah Wordsworth, Carla Wright, Sara Yadav, and Anna Zalevski. The team worked with the British Society for Antimicrobial Chemotherapy (BSAC) to deliver an elearning course that includes preliminary findings from the work in this manuscript. A payment was made to BSAC to help develop the course format and support platform administration. We would also like to acknowledge and thank Natasha Lal for additional editing support.

