## Supplemental file 1 for "How, why and when are delayed (back-up) antibiotic prescriptions used in primary care? A realist review integrating concepts of uncertainty in healthcare"

**Primary search strategy**

| Search stages | MeSH terms or equivalent in database |
|---|---|
| Delayed antibiotic prescribing | “delayed antibiotic*”<br>“delayed prescribing”<br>“back-up antibiotic*”<br>“back-up prescribing” |
| Combinations | (1 AND 3) OR (1 AND 2) OR 4 |
| Combination adjusted following input from librarian | ((delay* or back up or backup or back-up or just in case or (wait adj2 see)) adj2 prescri*) |
| The initial search terms were based on a previous systematic literature review around the effectiveness of delayed antibiotics for respiratory tract infections.[1] However, specific search terms related to the type of infection were excluded in our review. The term ‘back-up’ prescription was included to broaden the search and maximise the return of relevant documents. Back-up prescriptions (specifically antibiotics) pre-dates delayed prescriptions and are historically different in that the former is associated with providing a ‘just in case’ prescription for patients with long-term conditions and who are susceptible to repeat acute infections e.g. infective exacerbations of chronic obstructive pulmonary disease. Delayed prescriptions were later introduced as an intervention to ameliorate the use immediate antibiotics for any patients with an acute infection that may be of viral origin. However, recent studies suggest that the term delayed prescriptions can be misleading to patients and back-up prescriptions would be more acceptable, thus both terms may be used interchangeably by some clinicians and researchers. Our search therefore included both terms, and we also used a combination of free-text and relevant subject headings to the database being searched. | |
