## Supplemental file 2 for "How, why and when are delayed (back-up) antibiotic prescriptions used in primary care? A realist review integrating concepts of uncertainty in healthcare"

**Document characteristics: source, use, and nature of evidence**

| Data Source | Use | Nature of source/description |
|---|---|---|
| Literature | Formal data | 76 articles from Medline, EMBASE, Web of Science, Cochrane Database of Systematic Reviews, and PsycINFO. There were 28 articles from the UK; 15 from the USA; six from Australia; five from Ireland; four each from the Netherlands and New Zealand; three each from Norway, Canada and Spain; one each from Denmark, Ghana, Israel and Malta; one was undetermined. There were 50 empirical studies (including three quality improvement studies): 15 qualitative studies; 18 randomised trials; four other types of trials; nine observational; three quantitative; one mixed-methods. Twenty-three empirical studies were from the UK; six from the USA; five from Ireland; three each from Australia, Spain, and Norway; the remainder from New Zealand, the Netherlands, Italy, Canada, Denmark, Israel, Ghana, Malta. We included nine literature reviews and 17 commentary articles.<br><br>Included articles are listed alphabetically below. |
| Interviews | Formal data | Excerpts from 41 interviews conducted as part of wider 'STEP-UP' study (https://www.expmedndm.ox.ac.uk/step-up) to explore experiences of Clinical Commissioning Group and general practice professionals in implementing antimicrobial stewardship interventions.[1] Only excerpts focused on participants' discussion of DP use were included in this review. |
| Wider STEP-UP team and advisory group | Feedback, advice and sense-making | Discussion and feedback from four face-to-face meetings (three in the regular wider STEP-UP team meetings and one as part of the advisory group meeting). In addition to the multi-professional and interdisciplinary expertise of the wider STEP-UP team, the advisory panel had representation from a lay member, commissioner, national policy maker and an epidemiologist. |

### Included articles


Agnew, J. *et al.* (2013) "Delayed Prescribing of Antibiotics for Respiratory Tract Infections: Use of Information Leaflets," *Irish Medical Journal*, 106(8), pp. 243–4.

Alonso-Coello, P., Llor, C. and de la Poza Abad, M. (2016) "Mixed Diagnoses and Mixed Messages—Reply," *JAMA Internal Medicine*, 176(5).

Andrews, T. *et al.* (2012) "Interventions to influence consulting and antibiotic use for acute respiratory tract infections in children: A systematic review and Meta-Analysis," *PLoS ONE*, 7(1).

Arnold, S.R. and Straus, S.E. (2005) "Interventions to improve antibiotic prescribing practices in ambulatory care," *Cochrane Database of Systematic Reviews*, 15(16).

Arroll, B. *et al.* (2002) "Delayed antibiotic prescriptions: what are the experiences and attitudes of physicians and patients?," *Journal of Family Practice*, 51(11), pp. 954–959.

Arroll, B. *et al.* (2003) "Delayed prescriptions Can reduce antibiotic use in acute respiratory infections," *BMJ*, 327, pp. 1361–2.

Arroll, B., Kenealy, T. and Kerse, N. (2002) “Do Delayed Prescriptions Reduce the Use of Antibiotics for the Common Cold? A Single-Blind Controlled Trial,” *Journal of Family Practice*, 51(4), pp. 324–328.

Arroll, B., Kenealy, T. and Kerse, N. (2003) “Do delayed prescriptions reduce antibiotic use in respiratory tract infections? A systematic review ,” *British Journal General Practice*, 53(496), pp. 1–7.

Ashdown, H.F. *et al.* (2016) “Prescribing antibiotics to ‘at-risk’ children with influenza-like illness in primary care: Qualitative study,” *BMJ Open*, 6(6).

Boiko, O. *et al.* (2020) “Risks of use and non-use of antibiotics in primary care: qualitative study of prescribers’ views,” *BMJ Open*, 10(10), p. e038851.

Borek, A.J. *et al.* (2020) “Social and contextual influences on antibiotic prescribing and antimicrobial stewardship: A qualitative study with clinical commissioning group and general practice professionals,” *Antibiotics*, 9(12), pp. 1–15.

Borek, A.J. *et al.* (2021) “Implementing interventions to reduce antibiotic use: a qualitative study in high-prescribing practices,” *BMC Family Practice*, 22(1).

Bozzella, M.J. *et al.* (2018) “From paper to practice: Strategies for improving antibiotic stewardship in the pediatric ambulatory setting,” *Current Problems in Pediatric and Adolescent Health Care*, 48(11), pp. 289–305.

Cals, J.W.L. *et al.* (2010) “Point-of-care c-reactive protein testing and antibiotic prescribing for respiratory tract infections: A randomized controlled trial,” *Annals of Family Medicine*, 8(2), pp. 124–133.

Cates, C. (2003) “Delayed prescriptions in primary care,” *British Journal of General Practice*, 53(496), pp. 836–837.

Cimolai, N. (2020) “Delayed Antibiotic Prescribing in the Outpatient Setting,” *Journal of Pharmacy Practice*, 33(6), pp. 736–737.

Cooper, E. *et al.* (2020) “Diagnosis and Management of UTI in Primary Care Settings—A Qualitative Study to Inform a Diagnostic Quick Reference Tool for Women Under 65 Years,” *Antibiotics*, 9(9), p. 581.

Couchman, G.R., Rascoe, T.G. and Forjuoh, S.N. (2000) “Back-up Antibiotic Prescriptions for Common Respiratory Symptoms ,” *Journal of Family Practice*, 49(10).

Dallas, A. *et al.* (2020) “Delayed prescribing of antibiotics for acute respiratory infections by GP registrars: a qualitative study,” *Family Practice*, 37(3), pp. 406–411.

Damoiseaux, R. (2004) “Delayed prescriptions — not a good option for infants,” *The British Journal of General Practice*, 54(498), p. 58.

Dharod, A. (2016) “Delayed Prescriptions for Reducing Antibiotic Use ,” *Journal of Clinical Outcomes Management*, 23(3).

Dowell, J *et al.* (2001) *A randomised controlled trial of delayed antibiotic prescribing as a strategy for managing uncomplicated respiratory tract infection in primary care*.

Duane, S. *et al.* (2016) “Exploring experiences of delayed prescribing and symptomatic treatment for urinary tract infections among general practitioners and patients in ambulatory care: A qualitative study,” *Antibiotics*, 5(3).

Flynn, M. and Hooper, G. (2020) “Antimicrobial stewardship though FeverPAIN score: Successes and challenges in secondary care,” *Clinical Infection in Practice*, 7–8, p. 100024.

Francis, N.A. *et al.* (2012) “Delayed antibiotic prescribing and associated antibiotic consumption in adults with acute cough,” *British Journal of General Practice*, 62(602), pp. e639–e646.

Ghebrehewet, S. *et al.* (2020) “Implementation of a Delayed Prescribing Model to Reduce Antibiotic Prescribing for Suspected Upper Respiratory Tract Infections in a Hospital Outpatient Department, Ghana,” *Antibiotics*, 9(11), p. 773.

Hansen, P. (2016) *Antibiotic Resistance: Use of Delayed Prescriptions for Viral Syndromes in Urgent Care*. Available at: http://scholarship.shu.edu/final-projects/13.

Harris, D.J. (2013) “Initiatives to improve appropriate antibiotic prescribing in primary care,” *Journal of Antimicrobial Chemotherapy*, 68(11), pp. 2424–2427.

Hay, A.D. (2004) “Review: delaying a prescription reduces antibiotic use in upper respiratory tract infections,” *Evidence-Based Medicine*, 9(4).

Hay, A.D. *et al.* (2019) “Anaesthetic–analgesic ear drops to reduce antibiotic consumption in children with acute otitis media: the CEDAR RCT,” *Health Technology Assessment*, 23(34), pp. 1–48.

Hayes, M., Faherty, A. and Hannon, D. (2013) “Delayed prescriptions: attitudes and experiences of General Practitioners in the Midwest,” *Irish Medical Journal* [Preprint].

Høye, S., Frich, J.C. and Lindbæk, M. (2011) “Use and feasibility of delayed prescribing for respiratory tract infections: A questionnaire survey,” *BMC Family Practice*, 12(34).

Høye, S., Frich, J.C. and Lindbœk, M. (2010) “Delayed prescribing for upper respiratory tract infections: A qualitative study of GPs’ views and experiences,” *British Journal of General Practice*, 60(581), pp. 907–912.

Høye, S., Gjelstad, S. and Lindbæk, M. (2013) “Effects on antibiotic dispensing rates of interventions to promote delayed prescribing for respiratory tract infections in primary care,” *British Journal of General Practice*, 63(616), pp. e777–e786.

Hughes, A. *et al.* (2016) *Evaluating a point-of-care C-reactive protein test to support antibiotic prescribing decisions in a general practice-a-point-of-care-c-reactive-protein-test-to-support-antibiotic-prescribing-decisions-in-a*, *Clinical Pharmacist*.

Ivers, N., Arroll, B. and Allan, M. (2011) “Delayed antibiotic prescriptions for URTIs,” *Canadian Family Physician*, 57(11), p. 1287.

Johnson, J.R. (2007) “Wait-and-See Prescription for Acute Otitis Media,” *JAMA*, 297(2), pp. 152–153.

Kavanagh, K.E. *et al.* (2011) “A pilot study of the use of near-patient C-Reactive Protein testing in the treatment of adult respiratory tract infections in one Irish general practice,” *BMC Family Practice*, 12.

Lacy, S.M. (2007) “Writing a wait-and-see prescription for the treatment of acute otitis media may decrease the use of antibiotics,” *The Journal of Pediatrics*, 150(3), p. 319.

de La Poza Abad, M. *et al.* (2016) “Prescription strategies in acute uncomplicated respiratory infections a randomized clinical trial,” *JAMA Internal Medicine*, 176(1), pp. 21–29.

de La Poza Abad, M. *et al.* (2019) “Use of delayed antibiotic prescription in primary care: A cross-sectional study,” *BMC Family Practice*, 20(1).

Linder, J.A. and Friedberg, M.W. (2016) “Mixed Diagnoses and Mixed Messages,” *JAMA Internal Medicine*, 176(5), pp. 718–719.

Link, T.L. *et al.* (2016) “Reducing inappropriate antibiotic prescribing for adults with acute bronchitis in an urgent care setting a quality improvement initiative,” *Advanced Emergency Nursing Journal*, 38(4), pp. 327–335.

Little, P. *et al.* (1997) “Open randomised trial of prescribing strategies in managing sore throat,” *BMJ*, 314(722).

Little, P. (2001) “Pragmatic randomised controlled trial of two prescribing strategies for childhood acute otitis media,” *BMJ*, 322, pp. 336–342.

Little, P. (2005) “Delayed prescribing of antibiotics for upper respiratory tract infection,” *BMJ*, 331(301).

Little, P. *et al.* (2005) “Information Leaflet and Antibiotic Prescribing Strategies for Acute Lower Respiratory Tract Infection A Randomized Controlled Trial,” *JAMA*, 293(24), pp. 3029–3035.

Little, P. (2006) *Delayed Prescribing-A Sensible Approach to the Management of Acute Otitis Media*.

Little, P. *et al.* (2009) “Dipsticks and diagnostic algorithms in urinary tract infection: Development and validation, randomised trial, economic analysis, observational cohort and qualitative study,” *Health Technology Assessment*, 13(19).

Little, P. *et al.* (2010) “Effectiveness of five different approaches in management of urinary tract infection: Randomised controlled trial,” *BMJ (Online)*, 340, p. c199.

Little, P. *et al.* (2013) “Clinical score and rapid antigen detection test to guide antibiotic use for sore throats: Randomised controlled trial of PRISM (primary care streptococcal management),” *BMJ (Online)*, 347.

Little, P. *et al.* (2014) “Delayed antibiotic prescribing strategies for respiratory tract infections in primary care: Pragmatic, factorial, randomised controlled trial,” *BMJ (Online)*, 348, p. g1606.

Little, P. *et al.* (2017) “Antibiotic prescription strategies and adverse outcome for uncomplicated lower respiratory tract infections: Prospective cough complication cohort (3C) study,” *BMJ (Online)*, 357, p. j2418.

Little, P. (2020) “Delayed Antibiotic Prescriptions,” *JAMA*, 324(13), pp. 1352–1353.

Lum, E.P.M. *et al.* (2018) “Antibiotic prescribing in primary healthcare: Dominant factors and trade-offs in decision-making,” *Infection, Disease and Health*, 23(2), pp. 74–86.

McCullough, A.R. and Glasziou, P.P. (2016) “Delayed antibiotic prescribing strategies-time to implement?,” *JAMA Internal Medicine*. American Medical Association, pp. 29–30.

McDonagh, M. *et al.* (2016) *Improving Antibiotic Prescribing for Uncomplicated Acute Respiratory Tract Infections*.

Moore, M. *et al.* (2009) “Effect of antibiotic prescribing strategies and an information leaflet on longer-term reconsultation for acute lower respiratory tract infection,” *British Journal of General Practice*, 59(567), pp. 728–734.

Morrell, L. *et al.* (2020) “Delayed Antibiotic Prescription by General Practitioners in the UK: A Stated-Choice Study,” *Antibiotics*, 9(9).

Peters, S. *et al.* (2011) “Managing self-limiting respiratory tract infections: A qualitative study of the usefulness of the delayed prescribing strategy,” *British Journal of General Practice*, 61(590), pp. e579–e589.

Pshetizky, Y., Naimer, S. and Shvartzman, P. (2003) “Acute otitis media - A brief explanation to parents and antibiotic use,” *Family Practice*, 20(4), pp. 417–419.

Raft, C.F. *et al.* (2017) “Delayed antibiotic prescription for upper respiratory tract infections in children under primary care: Physicians’ views,” *European Journal of General Practice*, 23(1), pp. 191–196.

Redmond, N.M. *et al.* (2018) “Impact of antibiotics for children presenting to general practice with cough on adverse outcomes: Secondary analysis from a multicentre prospective cohort study,” *British Journal of General Practice*, 68(675), pp. e682–e693.

Rowe, T. and Linder, J.A. (2019) “Novel approaches to decrease inappropriate ambulatory antibiotic use,” *Expert Review of Anti-infective Therapy*, 17(7), pp. 511–521.

Rowe, T. and Linder, J.A. (2020a) “Delayed Antibiotic Prescriptions in Ambulatory Care: Reconsidering a Problematic Practice,” *JAMA - Journal of the American Medical Association*. American Medical Association, pp. 1779–1780.

Rowe, T. and Linder, J.A. (2020b) “Delayed Antibiotic Prescriptions—Reply,” *JAMA*, 324(13), p. 1353.

Ryves, R. *et al.* (2016) “Understanding the delayed prescribing of antibiotics for respiratory tract infection in primary care: A qualitative analysis,” *BMJ Open*, 6(11).

Saliba-Gustafsson, E.A. *et al.* (2019) “General practitioners’ perceptions of delayed antibiotic prescription for respiratory tract infections: A phenomenographic study,” *PLOS ONE*, 14(11), p. e0225506.

Sargent, L. *et al.* (2016) “Is Australia ready to implement delayed prescribing in primary care? A review of the evidence,” *Australian Family Physician*, 45(9), pp. 688–90.

Sargent, L. *et al.* (2017) “Using theory to explore facilitators and barriers to delayed prescribing in Australia: a qualitative study using the Theoretical Domains Framework and the Behaviour Change Wheel,” *BMC Family Practice*, 18(1).

Septimus, E.J. *et al.* (2017) “Extended-Release Guaifenesin/Pseudoephedrine Hydrochloride for Symptom Relief in Support of a Wait-and-See Approach for the Treatment of Acute Upper Respiratory Tract Infections: A Randomized, Double-Blind, Placebo-Controlled Study,” *Current Therapeutic Research - Clinical and Experimental*, 84, pp. 54–61.

Siegel, R.M. *et al.* (2003) “Treatment of Otitis Media With Observation and a Safety-Net Antibiotic Prescription,” *Pediatrics*, 112(3), pp. 527–531. Available at: www.aappublications.org/news.

Spiro, D.M. *et al.* (2006) *Wait-and-See Prescription for the Treatment of Acute Otitis Media A Randomized Controlled Trial*.

Spurling, G.K.P. *et al.* (2017) “Delayed antibiotic prescriptions for respiratory infections,” *Cochrane Database of Systematic Reviews* [Preprint]. John Wiley and Sons Ltd.

Thoolen, B., de Ridder, D. and van Lensvelt-Mulders, G. (2012) “Patient-oriented interventions to improve antibiotic prescribing practices in respiratory tract infections: A meta-analysis,” *Health Psychology Review*, 6(1), pp. 92–112.

Vellinga, A. *et al.* (2016) “Intervention to improve the quality of antimicrobial prescribing for urinary tract infection: A cluster randomized trial,” *CMAJ*, 188(2), pp. 108–115.

Vervloet, M. *et al.* (2016) “Reducing antibiotic prescriptions for respiratory tract infections in family practice: Results of a cluster randomized controlled trial evaluating a multifaceted peer-group-based intervention,” *npj Primary Care Respiratory Medicine*, 26.
