## Supplemental file 3 for "How, why and when are delayed (back-up) antibiotic prescriptions used in primary care? A realist review integrating concepts of uncertainty in healthcare"

**Categorising contexts, mechanisms and outcomes**

The concepts of context, mechanism and outcome can be challenging to define, even among realist experts, and the same thing can be a context in one circumstance, a mechanism in another, and an outcome in yet another—what is important is how these explain how and why something happens.(1) We therefore took a pragmatic approach to classifying contexts and mechanisms to produce four outcomes: immediate antibiotics; delayed prescription (DP) instead of immediate antibiotics; DP instead of no antibiotics; and no antibiotics. We identified 'contexts' and 'mechanisms' labels where the former were directly observable or perceived and the latter were mainly invisible, e.g. beliefs. Three researchers (AC, MMc, GW) organised these into potential context-mechanism-outcome configurations (CMOCs). We designated items as 'context' or 'mechanism' when we found evidence that their positions in the CMOC could be swapped/changed.(1)
