## Supplementary figures and images for "How, why and when are delayed (back-up) antibiotic prescriptions used in primary care? A realist review integrating concepts of uncertainty in healthcare"

### Supplemental file 4

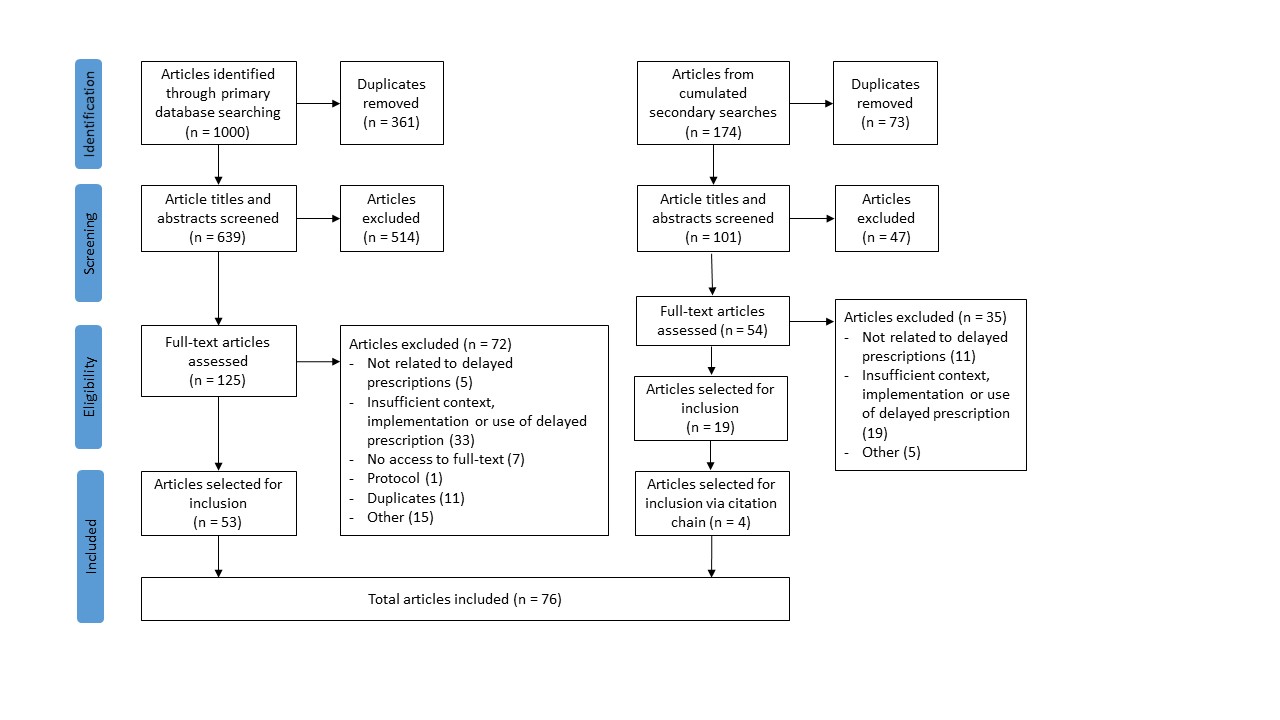

### Supplemental file 5

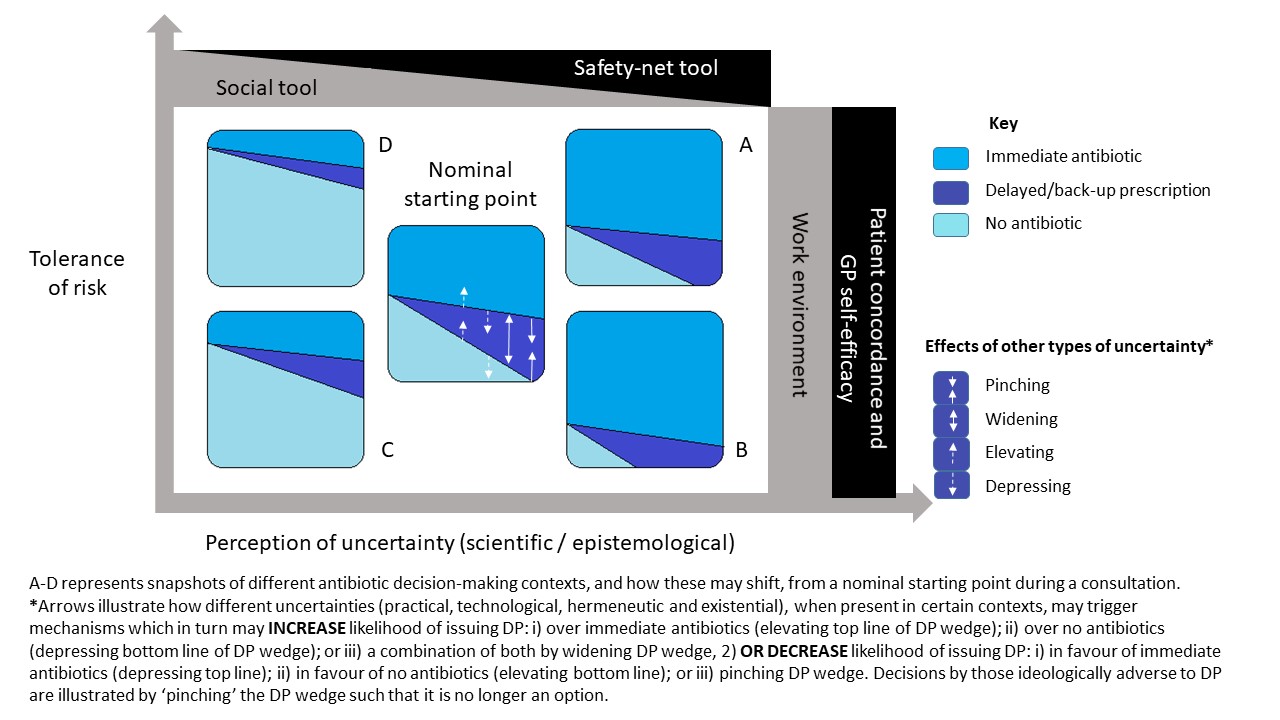
