## Supplemental file 6 for "How, why and when are delayed (back-up) antibiotic prescriptions used in primary care? A realist review integrating concepts of uncertainty in healthcare"

**Supplemental File 5**

**Summary of program theory key CMOC topics and associated uncertainty types with example supporting data**

| **Program theory key CMOC topics** | **Uncertainty type*** | **Example supporting data** |
|---|---|---|
| **I. The value of DP to safety-net** | | |
| • <u>Key CMOC 1</u> Antibiotic access<br><br>• <u>Key CMOC 2</u> Patients at high risk of developing complications | **Epistemological**<br>**Scientific**<br><br>DP use was influenced by how GPs' epistemological/scientific uncertainty was addressed when they interpret guidelines as either supporting DP use or cautioning against it. | "We found that GPs issue wait-and-see prescription most commonly in sinusitis and otitis. When compared to a similar group of GPs in Norway… our numbers show an over-representation of sinusitis and otitis, which indicates that patients receiving antibiotics for otitis or sinusitis more often will be instructed to wait than patients receiving antibiotics for other conditions. This may be because otitis and sinusitis are the two conditions for which the Norwegian National Treatment Guidelines recommend "watchful waiting"." [1] |
| | **Existential**<br>**Scientific**<br><br>DP as a cause of existential and scientific uncertainty leading to beliefs that it is unprofessional or unsafe to use DP. | "Ultimately, delayed antibiotic prescriptions expose patients to unnecessary and potentially harmful medications and do not improve clinical outcomes. The appropriate course of action for most patients is providing reassurance that antibiotics are not indicated, providing clear instructions about symptomatic management, and communicating specific instructions for when patients should call or return if symptoms worsen or change. Health care professionals have a responsibility to do what is in the best interest of patients even if there are concerns that doing so will make some patients unhappy or unsatisfied. For patients with viral conditions, that responsibility means not prescribing immediate or delayed antibiotics."[2] |
| | **Hermeneutic**<br><br>DP may or may not help mitigate hermeneutic uncertainty around interpreting patient risk factors. | "Most physicians restricted delayed prescriptions to a particular age range. However, within this category there was considerable variability and inconsistency. Many used delayed prescriptions only for children, with children younger than 6 years being the most suitable group; others used delayed prescribing only for children older than 6 to 8 years. One physician would not use the strategy in very young children, i.e., younger than 3 years.[3] |
| | **Practical**<br>**Existential**<br><br>DP may help mitigate practical uncertainty about when patients can access antibiotic or reconsult and existential uncertainty about potential negative consequences on their relationship with patients. | "… it's a weekend, the long weekend, they're about to go away or they're travelling on a cruise or something, so they might be reasons why I would give them a script to use if needed."[4]<br><br>"We know people don't want to come to us twice because they don't want to pay the fee. . . So. . . we tell them. . . don't give your kid an antibiotic or try to avoid the antibiotic. . . I will give you a prescription. . . dated not for today. . . for two days' time, and then it's only valid for a week. . . if you get worse buy it. In that way, the patient doesn't think that we're not giving him an antibiotic so that he comes back to us again to pay us."[5] |
| **II. GP self-efficacy and patient concordance** | | |
| | **Epistemological**<br>**Scientific** | "Most GPs believed that it would be difficult to implement delayed prescribing: they do not trust patients to delay and |

| | | |
|---|---|---|
| • <u>Key CMOC</u> Patient concordance<br><br>• <u>Key CMOC 4</u> Feedback about DP use<br><br>• <u>Key CMOC 5</u> GP self-efficacy to explain DP rationale and instructions | Epistemological and scientific uncertainty around patient concordance with DP leading to doubts about DP benefits. These uncertainties may be mitigated with feedback and/or recontact about DP use. | do not believe there is enough evidence to refute this perception. (…) "Some people are just going to go out and fill the prescription anyway." (GP19)[6]<br><br>"I would also always insist even if they think they need it [the delayed antibiotic prescription], I would ask them to phone me, to tell me what's happening. At least I would have feedback and then I can make sure that they really need it."[5] |
| | **Hermeneutic**<br><br>DP was perceived to be a challenge when there was hermeneutic uncertainty around interpreting a patient's ability to use DP appropriately. | "Physicians generally were selective about patients for whom they considered delayed prescribing appropriate. Patients who were poorly educated, who had a bad command of English, or who were transient to the practice were identified as poor candidates for receiving delayed prescriptions." [3] |
| | **Existential**<br><br>DP as a cause of existential uncertainty around how the clinician might be perceived by patients. | "But others felt it could damage relationships, as patients may perceive delayed prescribing as an indication of clinician incompetence or a way of saving money."[7] |
| | **Practical**<br><br>DP can contribute or mitigate practical uncertainty around how patients will use the antibiotic. | "… the patient should receive thorough information and clear instructions about when to start the medication. This was to ensure that the patient understood that there was a proper judgement behind the use of delayed prescribing, and to prevent patients from demanding a wait-and-see prescription for any respiratory tract infection. One GP said: 'If we just deliver a prescription and say "Start with the medication if you're not better in 4 days", without giving more information about the condition and explain a little, then I think [the strategy] might have a slippery-slope effect. There needs to be a conversation to prevent this from happening'." [1]<br><br>"Those GPs with higher levels of experience expressed higher perceptions of self-efficacy in using delayed prescribing strategies: As time goes by I think you develop better explanations for patients that mean when you are explaining to them why they don't need an antibiotic you are more convincing." (GP31, low prescribing)[8] |
| | **Practical**<br>**Technological**<br><br>Practical/technological uncertainty around how different DP formats can be used effectively. | "However, some GPs from high-prescribing practices found delayed prescribing strategies were complex and the uncertainty in administering them led to difficulties in successfully delaying a prescription. These difficulties included being able to fully understand the method of delayed prescribing and the confidence in using the strategy during the consultation, particularly when met with perceived patient pressure for an antibiotic prescription."[8] |
| **III. Social tool value of DP** | | |

| | | |
|---|---|---|
| • <u>Key CMOC 6</u> Minimising conflict<br><br>• <u>Key CMOC 7</u> Patient education & empowerment | **Epistemological**<br>**Scientific**<br><br>Epistemological/scientific uncertainty around how use of antibiotics (including DP) influence patient expectations for antibiotics and subsequent impact on antibiotic use and resistance. | “Furthermore, the effect of prescribing antibiotics on belief in and expectation for antibiotics must be quantified: the cycle of prescribing and expectation is likely to encourage attendance in future episodes, increase pressure on doctors to prescribe, increase antibiotic use, and increase the danger of antibiotic resistance.”[9] |
| | **Existential**<br><br>DP may help mitigate or contribute to existential uncertainty about a GP’s role to balance professional responsibility with empowering patients. | “I like this approach– it [delayed prescribing] gives the patient some control in their sickness, isn’t this what we want patients to do? Surely this is a good thing, helping patients take more responsibility and educating them.” (GP19) [6]<br><br>“Well, I would say that it’s probably often used by people as a bit of a get-out-of-jail-free card ….I feel like that’s sort of failing a little bit in that if they’re not meant to have antibiotics, you haven’t given really a very clear answer there and a clear message to the patient.”[4] |
| | **Hermeneutic**<br><br>DP may help mitigate hermeneutic uncertainty around interpreting symptom severity for those unable to express themselves clearly. | “Most commonly, a delayed prescription was given for symptoms suggestive of upper respiratory tract infections. “Generally it’s a parent of a child that feels that the child needs an antibiotic but the child seems reasonably well and all parameters are within normal limits. It’s probably a learnt response from a previous inappropriate prescription.” (Young Female rural GP) [10] |
| | **Practical**<br>**Existential**<br><br>DP may help mitigate practical uncertainty for how to end a consultation and existential uncertainty around the potential impact of not addressing patient expectations. | “One GP explained that delayed prescribing offered a compromise when the GP was of the opinion that antibiotics were not appropriate but the patient desired an antibiotic prescription. If GPs were encountering difficulties in ending a consultation then a delayed prescription was described as a closing strategy: ‘I think it’s often used in a situation where the clinician doesn’t think antibiotics are justified, but the patient is adamant that they would like them and it’s seen as a compromise. So it’s basically a way to end the consultation where both parties can feel happy that they’ve got what they wanted’.” (GP31, high prescribing)[8]<br><br>“Although some prescribers saw RTI consultations as being relatively simple, they were viewed by others as challenging, due to patients’ expectations of antibiotics. These expectations were believed to have arisen from patients’ previous experiences of receiving an antibiotic for an RTI and the perceived success of this treatment. ‘If you are dishing out an antibiotic that is an easy consultation. Whereas if you try and educate them that is where the difficulty starts because they have got that thing in their mind that whenever they have got a cough they have to [have an antibiotic].’ (GP9) ‘It’s difficult because they go |

| | | |
|---|---|---|
| | | "well last time I had this exact thing and Dr so and so gave me antibiotics, why won't you?"... it puts you in such a difficult [position] ... and sometimes I have had to sort of give delayed scripts even though I really don't want to'." (trainee GP8)[11] |
| **IV. Work environment** | | |
| • <u>Key CMOC 8</u> Workload and time pressures<br><br>• <u>Key CMOC 9</u> Practice culture and structure | **Practical**<br><br>DP may help mitigate and contribute practical uncertainty around how to balance workload with risk | "The physicians also emphasized a substantial workload as a reason for making them inclined to use delayed prescriptions, due to lack of time to follow up on the child"[12]<br><br>"However, some GPs found particular methods of delayed prescribing to be 'mutually inconvenient' (GP13, low prescriber) for the GP and the patient. One GP thought that if the method of telephone reconsultations was used, this created more work for the patient and the GP."[8] |
| | **Practical**<br>**Technological**<br>**Scientific**<br>**Epistemological**<br><br>There are practical and technological uncertainty around how DP use can be captured which contributes to lack of data/knowledge (scientific/ epistemological uncertainty) about its use, potential benefits and risks. | "That's [tracking DP] where the problem is. In EMIS there is a code for delayed prescriptions. The problem with coding is a huge issue. You try to look at coding for pneumonia. I can give you probably five things that it's been coded as. Coding is a huge issue and until we improve that, it's going to be really difficult to look at. That's why I ask them to leave a note in the consultation notes, so if I did go back to look at particular antibiotics that had been prescribed, then if it was a delayed prescription, then I could find out whether it had been dispensed or not or what is going on. At the moment, I don't actually have any robust way in picking out all the delayed prescriptions because of the coding issues and fiascos that I go through."[13] |
| | **Practical**<br>**Scientific**<br>**Epistemological**<br><br>Practical, scientific and epistemological uncertainty around how DP can be used effectively in practice. | "Despite a mixed response from GPs with regard to training in how to use delayed prescribing strategies effectively, there was some consensus of the need of a standardised policy between and within practices and [Clinical Commissioning Groups] to ensure that strategies are consistent throughout the country."[8] |
| ***Scientific** (around data relating to diagnosis, prognosis, cause and treatment); **Epistemological** (knowledge); **Practical** (structures/processes of care); **Technological** (skills/equipment/software); **Hermeneutic** (interpreting data and patients' narratives); **Existential** (around personal identity, worldview, sense of meaning) | | |
